# Functional mapping of B-cell linear epitopes of SARS-CoV-2 in COVID-19 convalescent population

**DOI:** 10.1101/2020.07.25.20161869

**Authors:** Zhigang Yi, Yun Ling, Xiaonan Zhang, Jieliang Chen, Kongying Hu, Yuyan Wang, Wuhui song, Tianlei Ying, Rong Zhang, Hongzou Lu, Zhenghong Yuan

## Abstract

Pandemic SARS-CoV-2 has infected over 10 million people and caused over 500,000 mortalities. Vaccine development is in urgent need to stop the pandemic. Despite great progresses on SARS-CoV-2 vaccine development, the efficacy of the vaccines remains to be determined. Deciphering the interactions of the viral epitopes with their elicited neutralizing antibodies in the convalescent COVID-19 population inspires the vaccine development. In this study, we devised a peptide array composed of 20-mer overlapped peptides of spike (S), membrane (M) and envelope (E) proteins, and performed a screening with 120 COVID-19 convalescent serums and 24 non-COVID-19 serums. We identified five SARS-CoV-2-specific dominant epitopes that reacted with above 40% COVID-19 convalescent serums. Epitopes in the receptor-binding domain (RBD) of S ill reacted with the convalescent serums. Of note, two peptides non-specifically interacted with most of the non-COVID-19 serums. Neutralization assay indicated that only five serums completely blocked viral infection at the dilution of 1:200. By using a peptide-compete neutralizing assay, we found that three dominant epitopes partially competed the neutralization activity of several convalescent serums, suggesting antibodies elicited by these epitopes played an important role in neutralizing viral infection. The epitopes we identified in this study may serve as vaccine candidates to elicit neutralizing antibodies in most vaccinated people or specific antigens for SARS-CoV-2 diagnosis.

## Introduction

Since the outbreak of COVID-19 in Wuhan China in December 2019, it was endemic to all over the China within less than one month and now become pandemic in over 200 countries. The infected cases surged to over 10 million by the end of June 2020, with over 500,000 mortalities (https://www.who.int/emergencies/diseases/novel-coronavirus-2019). COVID-19 is caused by infection with a novel coronavirus SARS-CoV-2 (1-3). Efficacy of several antiviral agents such as remdesivir and chloroquine remains uncertainty (4-6). Vaccine development is in urgent need to stop the pandemic. Coronaviruses contains three structural proteins on the surface of virus membrane envelope: the spike (S), membrane (M) and envelop (E) proteins. S protrudes from the virion surface to form a spike and interacts with host receptor to mediate virus entry (7). The spike protein (S) of SARS-CoV-2 interacts with host receptor ACE2 (2). SARS-CoV-2 vaccine candidates include viral vector expressing SARS-CoV-2 spike protein (S) (8), the receptor binding domain (RBD) (9) and inactivated virion (10). With the start of the first clinical trial of a recombinant adenovirus type-5 (Ad5) vectored COVID-19 vaccine (8), there would be many other vaccine trials. Despite great progresses on SARS-CoV-2 vaccine development, there are still critical questions remains to be answered, such as which kind of antigens (epitopes) elicit robust neutralizing antibodies in most people; if the convalescent serum of COVID-19 patients contains neutralizing antibodies and which viral epitopes are recognized by the neutralizing antibodies. Answers to these questions may guide development of effective vaccines. In this study, we collected the serums of a convalescent COVID-19 cohort. By using a peptide array composed of 20-mer overlapped peptides of S, M and E, we identified several dominant SARS-CoV-2 specific linear epitopes that reacted with the convalescent serums. By using peptide-compete neutralizing assay, we found that some of these dominant epitopes partially competed the neutralization activity of several convalescent serums. These data provide information for developing novel vaccine candidates and antigens for specific antibody diagnosis.

## Results

### Identification of dominant linear B-cell epitopes of COVID-19 by a peptide array

To map the dominant linear B-cell epitopes recognized by most COVID-19 patients, we designed a 20-mer overlapped peptide array encompassing the entire spike (S), the ectodomain of membrane (M), the loops between the transmembrane helices (TMs) and representative regions of endomains of M and envelope (E) proteins (Fig.1a and b)(Supplementary table 1). We used the peptide array to perform a screening with convalescent serums from 120 COVID-19 patients and 24 serums collected from patients with symptoms of acute respiratory infections but with negative diagnosis of COVID-19. The serums were diluted 1:200 and incubated with the peptide array, followed by incubated with HRP-conjugated anti-human IgG. Peptides reacting with the serums were visualized by chemiluminescent reagents. Of the 120 COVID-19 convalescent serums, 2 serums didn’t react with any peptide; 4 serums only reacted with one peptide in the peptide array. Peptides S1-52, S1-55, S1-57, S2-13, S2-47, M1 reacted with more than 40% COVID-19 serums (Fig. 1c and 1d). Of note, S1-52 and S2-56 reacted with more than 40% COVID-19-negative control serums, indicating non-specific reaction (Fig. 1c and 1d). We also detected about 17% serum reacted with a peptide (E2) in the endodomain of E whereas no reaction with the peptide in the ectodomain (E1) (Fig. 1c and 1d). The topology of E protein remains controversial. Evidences indicate the N-terminal part is the ectodomain (Fig. 1a)(11, 12). If the interaction of E endodomain with antibodies is due to the real topologically exposure remains to be clarified.

**Figure 1.**
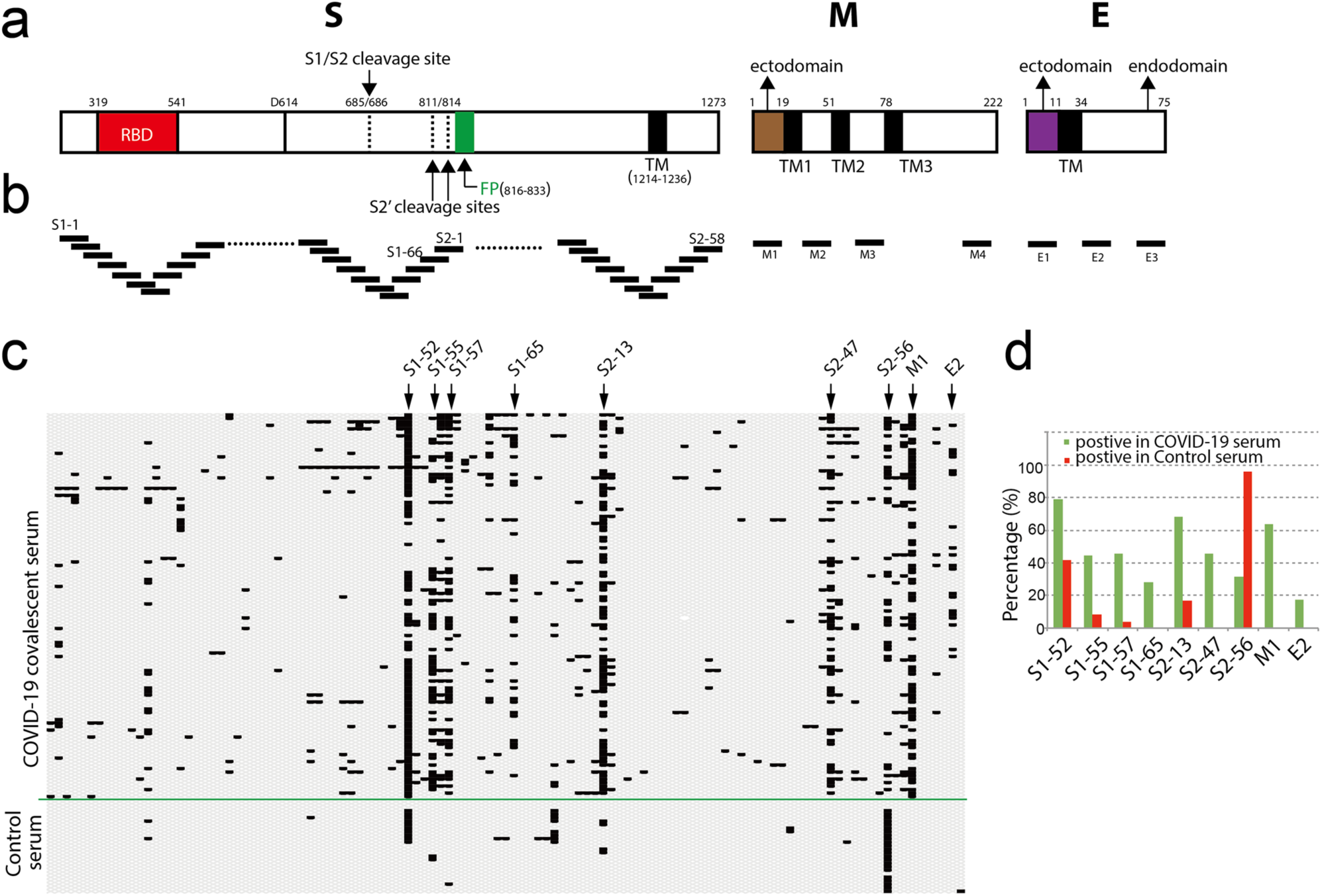
Screening convalescent serum of COVID-19 by peptide array. (a) Schematic of SARS-CoV-2 virion structural proteins: spike (S), membrane (M) and envelope (E) proteins. In the diagram of S, the receptor-binding domain (RBD) is in *red*. Fusion peptide (FP) is in *green*. Black boxes indicate transmembrane helices (TM). Arrows indicate S1/S2 cleavage site and alternative S2′ cleavages sites, respectively. In the diagrams of M and E, the endodomians and ectodomains are indicated. The numbers above the diagrams indicate the positions (aa) of each domain. (b) Design of peptide array. 20-mer overlapped peptides covering the entire S, the ectodomain of M, the loops between the TMs and representative regions of endomains of M and E were synthesized and doted. (c-d) Convalescent serums from COVID-19 patients and control serums from non-COVID-19 patients (control serum) were diluted (1:200) and incubated with peptide array, followed by incubated with HRP-conjugated anti-human IgG. Peptide array were visualized by chemiluminescent reagents, scanned and quantified. The peptides with signal to noise above 2 were counted as positive (black bar). The positive peptides for each patient were plotted. Arrows indicate representative peptides. (d) The positive percentage of the indicated peptides.

The peptides S1-52, S1-55, S1-57, S2-13, S2-47, M1 that reacted with more than 40% COVID-19 serums represented as dominant linear B-cell epitopes. We located these epitopes in a crystal structure of S in open sate (PBD, 6vyb) (Fig. 2a and 2b). The peptides S1-52 and S1-55 were near the receptor-binding domain (RBD) (Fig. 2a and 2b). The peptide S2-13 contained the alternative S2′ cleavages sites and fusion peptide (FP) (Fig. 2a and 2b). Most of the peptide S2-47 is not available in the crystal structure (Fig. 2a and 2b). Peptide S2-47 is conserved in SARS-CoV-2 and SARS-CoV whereas other peptide varied in SARS-CoV-2 and SARS-CoV (Fig. 2c).

**Figure 2.**
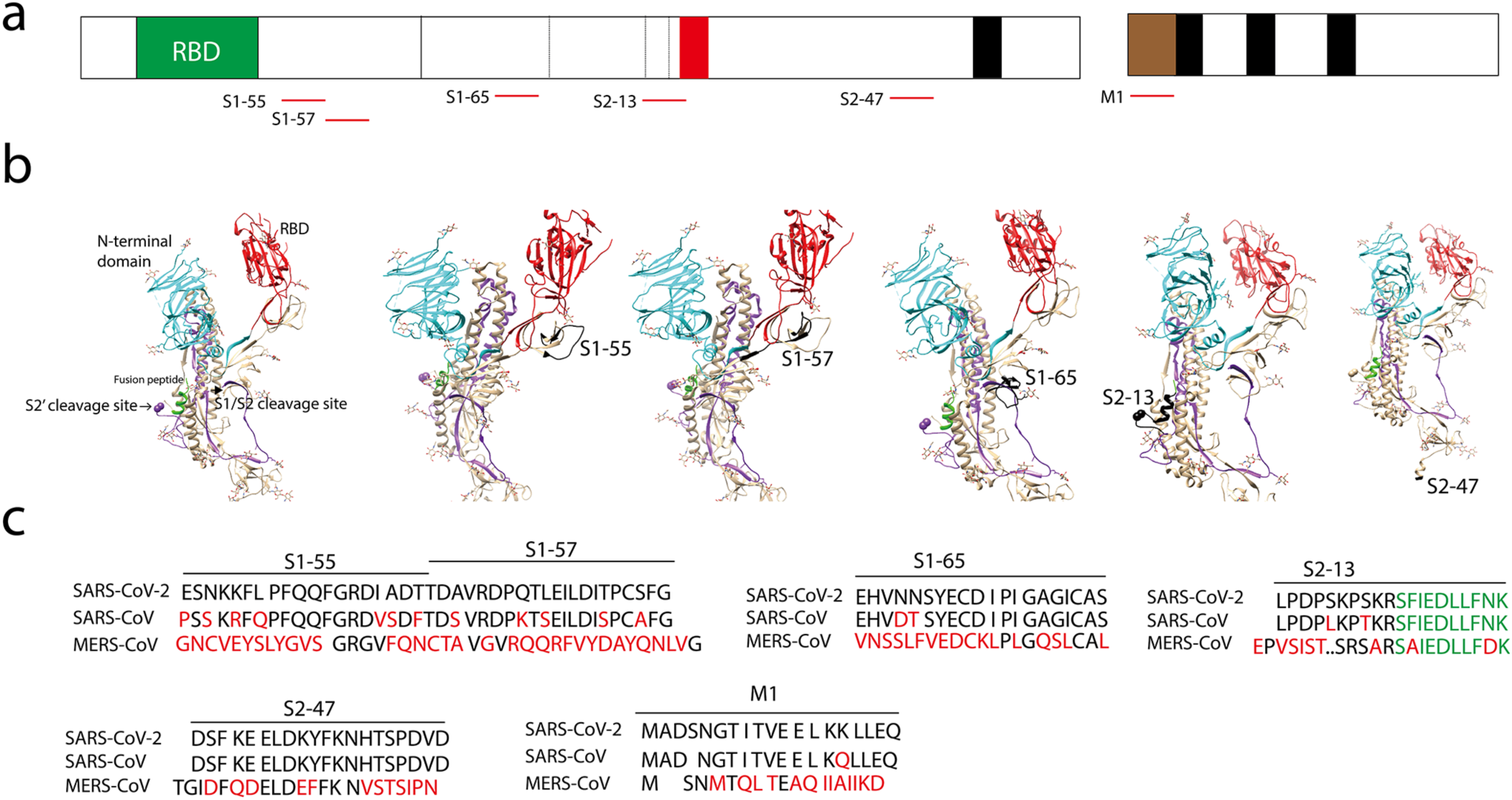
Identification of dominant B-cell linear epitopes of SARS-CoV-2 virus particle. (a) Schematic of spike (S), membrane (M) and the dominant B-cell linear epitopes (red lines). (b) Locations of the dominant B-cell linear epitopes in the S structure. The S structure (open sate; PBD, 6vyb), N-terminal domain (cyan), RBD (red), fusion peptide (green). The B-cell linear epitopes are in black. Only part of the S2-47 is visible in the structure. (c) Alignment of the dominant B-cell linear epitopes. Different amino acids are in red. Fusion peptide in S2-13 is in green.

### Less B-cell linear epitopes on Receptor binding domain (RBD) of Spike protein (S)

Receptor binding domain (RBD) of coronaviruses spike proteins interacts with host receptor to mediate virus entry and some antibodies against RBD potently neutralize viral infection (7). We examined the reactions of the RBD peptides with the serums (Fig. 3a). Except for S1-52, almost all the RBD peptides reacted with less than 10% serums, with peptides such as S1-31, S1-32, S1-34 and S1-35 that didn’t react with any serum (Fig. 3b). Although the peptide S1-52 interacted with most serums, it also non-specifically interacted with control serums (Fig. 3c). A poor-interaction peptide S1-31 was partially masked by the high-reaction peptide S1-52 in the crystal structures (Fig. 3c), suggesting the poor reaction of this peptide was probably due to its poor accessibility. In contrast, the peptide S1-32 and the ACE-2 interacting-interface (Fig. 3a, in red) have good accessibility (Fig. 3c) but still exhibited poor reactions (Fig. 3b), suggesting poor reactions of peptides in RBD in general.

**Figure 3.**
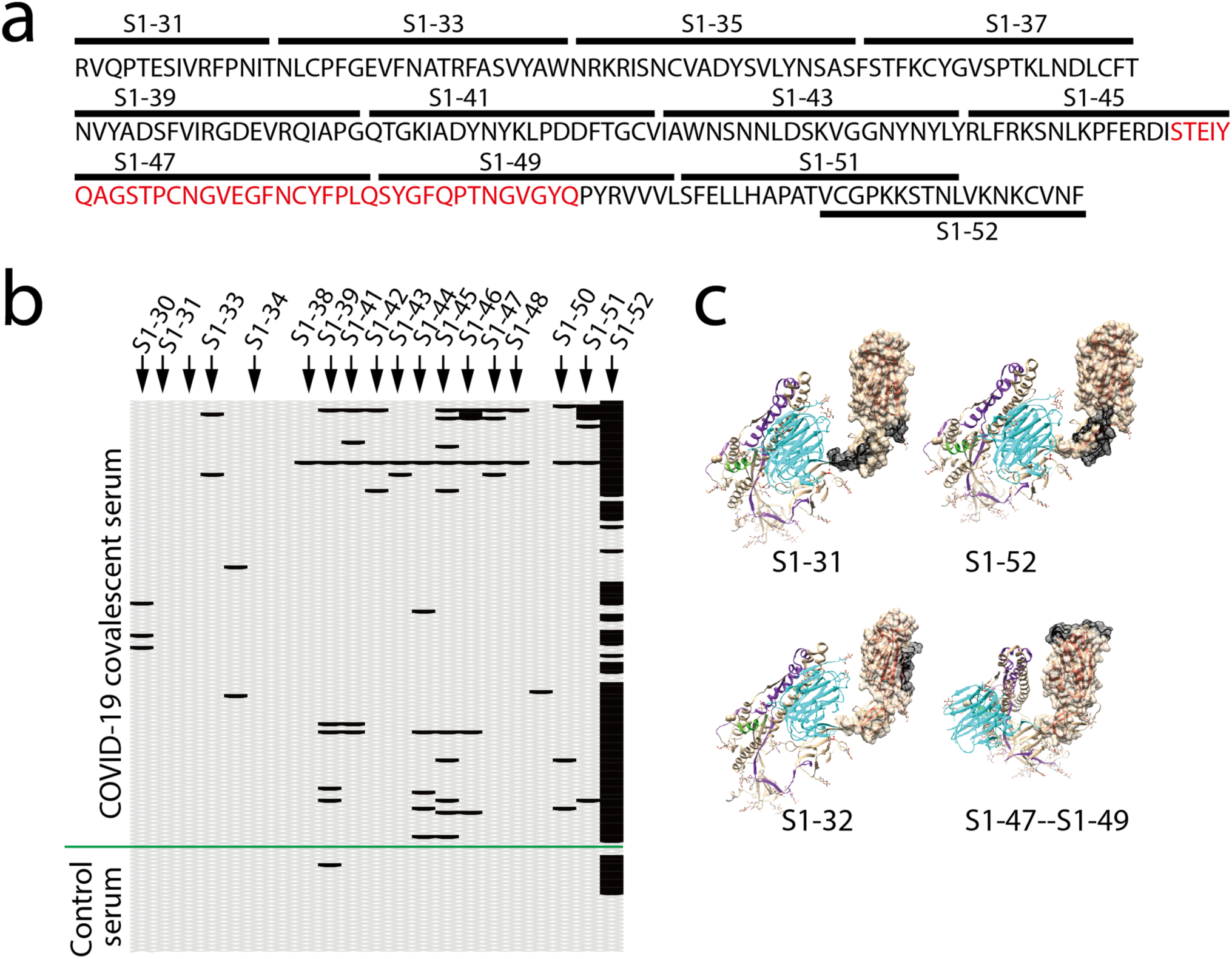
Less B-cell linear epitopes on Receptor binding domain (RBD) of Spike protein (S). (a) Amino acid sequence of RBD. Black bars indicate the corresponding peptides. The ACE-2 interacting interface was in red. (b) Reaction of COVID-19 convalescent serums with the peptides in RBD. The peptides with signal to noise above 2 were counted as positive (black bar). (c) Representative peptide (in black) in the structure of RBD (PBD, 6vyb).

### Neutralizing SARS-CoV-2 infection by COVID-19 convalescent serum

To explore the functional relevance of the identified B-cell linear epitopes and their elicited antibodies with the neutralizing activity, we first assessed the activities of the serums for the neutralization of SARS-CoV-2 virus infection. COVID-19 convalescent serums were diluted at 1:200 and incubated with 100 PFU (plague-forming unit) of virus. The neutralizing activity was assessed by reduction of viral infection-induced cytopathic effect (CPE) and quantification of viral RNA levels in the supernatants of the infected cells (Fig. 4a). We used a SARS-CoV-2 strain nCoV-SH01 that we isolated from the throat swab sample of a COVID-19 patient (13). Virus infection induces typical syncytial cytopathic effects on Vero E6 cells (supplementary Fig. 1). Some COVID-19 convalescent serums could partially reduce (CPE+, CPE++) the CPE or completely blocked the appearance of CPE (CPE-) (supplementary Fig. 1; Supplementary table 2). Of the 120 COVID-19 convalescent serums tested, at the dilution of 1:200, five serums completely blocked the appearance of CPE at 3 days post infection. And seven serums partially reduced the CPE or completely blocked the appearance of CPE in one of the duplicated wells (Supplementary table 2). We also quantified the viral RNA levels in the supernatants of the infected cells at 3 days post infection with some representative samples. Serums P13, P18, P68, P98 and P107 that completely blocked the appearance of CPE also dramatically reduced the viral RNA levels (Fig. 4b).

**Figure 4.**
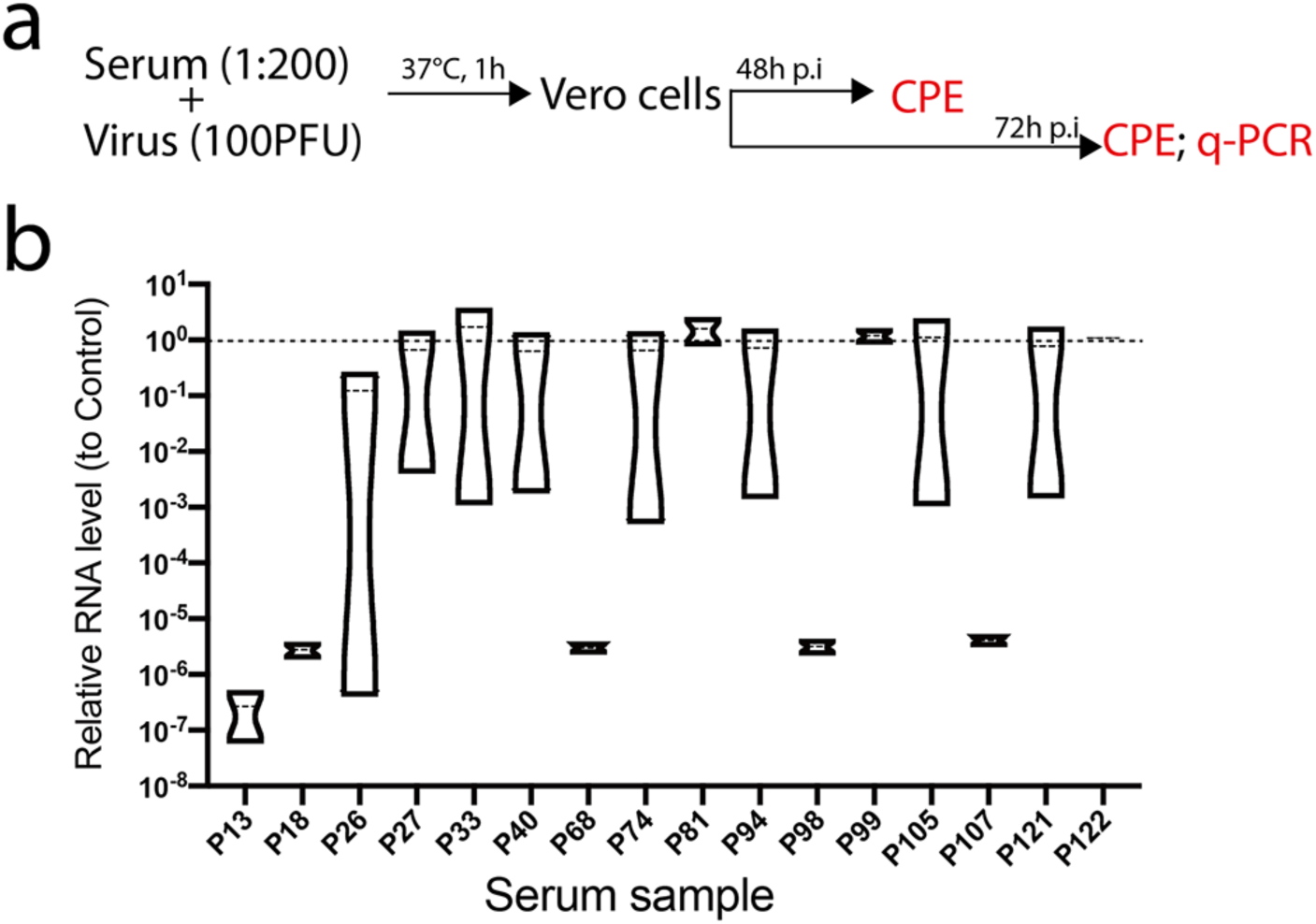
Neutralizing SARS-CoV-2 infection by COVID-19 convalescent serum. (a) Schematic of experiment design. COVID-19 convalescent serums were diluted and incubated with SARS-CoV-2 before added to the media. At 48 hours or 72 hours post infection (p.i), the cytopathic effect (CPE) of the infected cells was examined. The viral RNAs in the supernatants were quantified by real-time PCR (q-PCR). (b) Quantification of the viral RNA levels in the supernatants from representative wells. Relative RNA levels to the mock-treated wells were plotted (n=2).

### Competing the neutralizing activity of convalescent serum with dominant B-cell linear epitopes

Then we compared the peptide array reaction patterns of the neutralizing serums (Nab) with the patterns of several non-neutralizing serums (Non-Nab). All the Nabs reacted with the dominant epitopes S1-55, S1-57 and S2-13. In contrast, only part of the Non-nabs reacted with these epitopes (Fig. 5a). Next, we evaluated if the antibodies against these epitopes play a role in activity of neutralizing viral infection. We performed a neutralizing experiment by using the peptide S1-55, S1-57, S2-13 and S2-47 compete the neutralizing activity of four neutralizing serums (Fig. 5b). The peptides were added at a final concentration of to 0.25mg/ml, at which, typically the peptide could completely block the corresponding antibodies (14). Adding of the peptides alone didn’t affect viral infection. Out of the four serums tested, peptides S1-55, S1-57 and S2-47 partially competed the neutralizing activity of one or two of the serums (Fig. 5c), suggesting that antibodies against these peptides partially contribute to the neutralizing activity of the serums.

**Figure 5.**
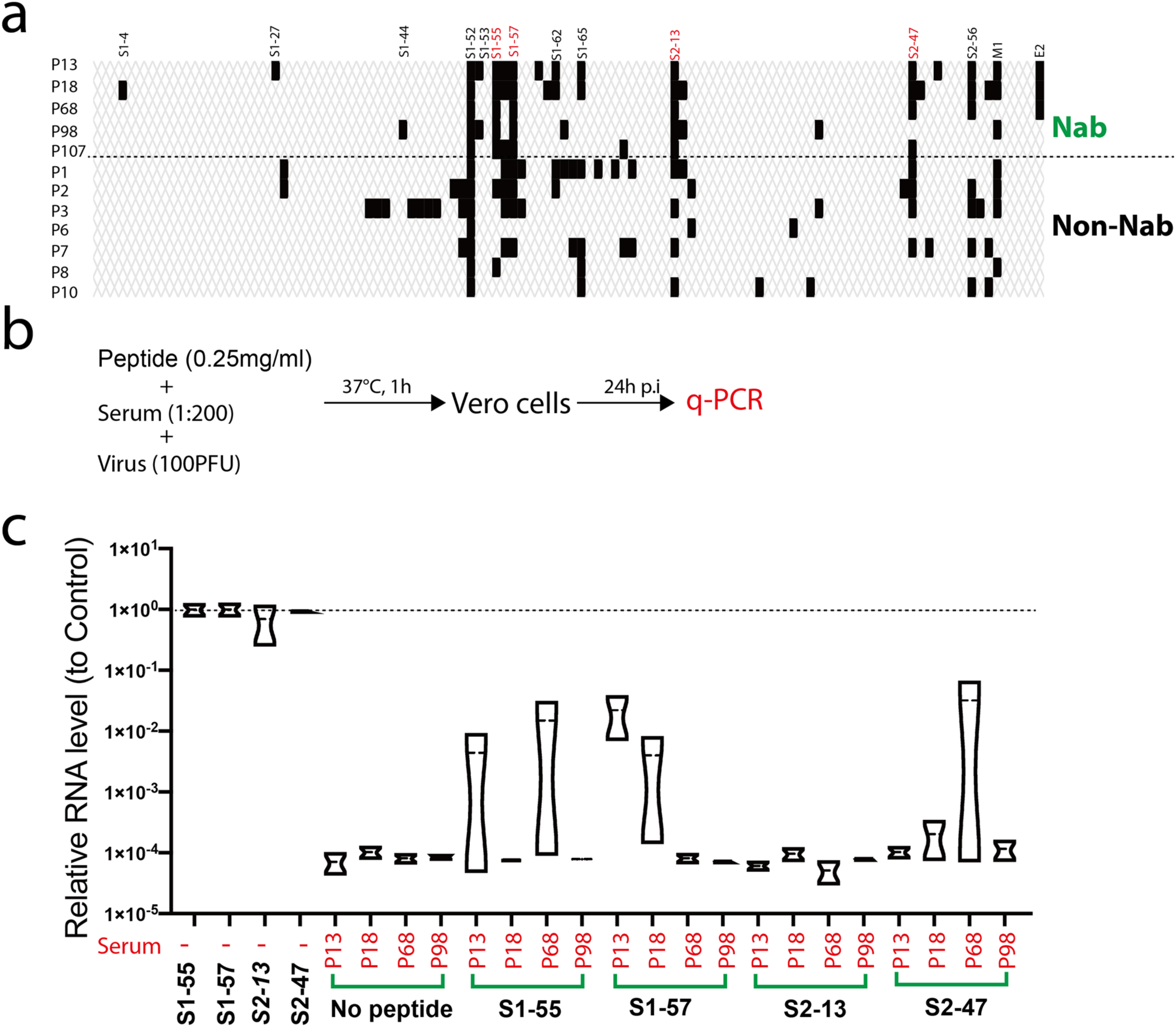
Competing the neutralizing activity of convalescent serum with dominant B-cell linear epitopes. (a) Reaction of the neutralizing antibodies (Nab) and non-neutralizing antibodies (Non-Nab) with the peptide array. The peptides with signal to noise above 2.1 were counted as positive (black bar). (b) Schematic of experiment design. Four neutralizing serums (P13, P18, P68 and P98) were diluted and incubated with SARS-CoV-2 and the peptides (S1-55, S1-57, S2-13 and S2-47), respectively before added to the media. At 24 p.i, the viral RNAs in the supernatants were quantified by q-PCR. Experiment was done in duplicated well for each serum (b) Quantification of the viral RNA levels in the supernatants from representative wells. Relative RNA levels to the mock-treated wells were plotted (n=2).

### Relationship of the neutralizing activity of the serums with the progress of the COVID-19 disease

To explore if the relationship of the neutralizing activity of the serums with the progress of the COVID-19 disease, we retrospectively analyzed the disease severity of the 120 COVID-19 patients. Eight patients experienced severe symptoms and patients who have serums with complete or partial neutralizing activity experienced asymptomatic or mild diseases (data not shown). We then examined the time intervals from the symptom onset to convalescence with SARS-CoV-2 triple negative diagnosis for throat swab, urine and feces. The average time interval for convalescence is 16.3 days. Patients with complete or neutralizing-antibodies (Fig. 6, in red and arrows) had variable time interval for convalescence, ranging from 8 days to 35 days, which was similar with the patients without neutralizing-antibodies (Fig. 6).

**Figure 6.**
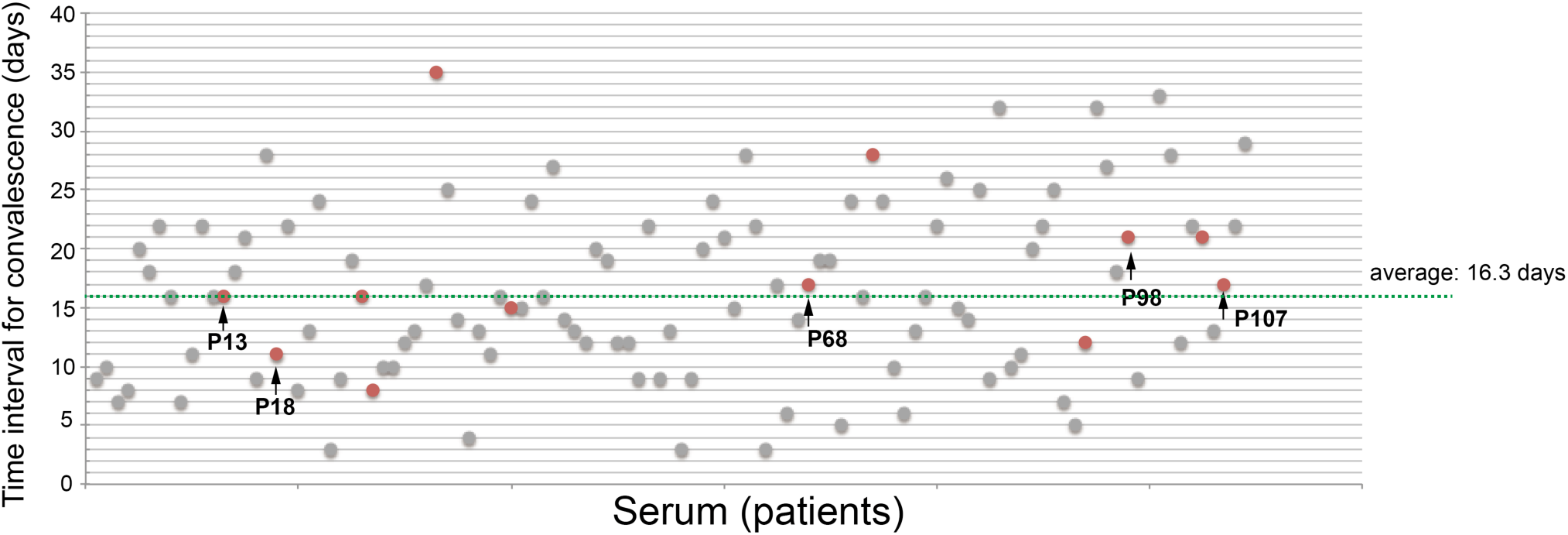
Time interval for convalescence. Time intervals from symptom onset to SARS-CoV-2 triple negative diagnosis for throat swab, urine and feces. Serums with neutralizing activity are in red. Arrows indicate fully neutralizing antibodies.

## Discussion

To learn the profile of the B-cell linear epitopes recognized by COVID-19 cohort for guiding vaccine design and devising diagnosis reagents, we performed functional B-cell linear epitope mapping with the convalescent serums of a COVID-19 cohort. By using a 20-mer-peptide array covering the S, M and E, we identified SARS-CoV-2 dominant epitopes recognized by most COVID-19 convalescent serums (Fig. 1 and 2). By using peptide-compete neutralizing assay, we found that epitopes in the peptides S1-55, S1-57 and S2-47 partially competed the neutralization activity of several convalescent serums (Fig. 5), suggesting antibodies against these epitopes play an important role in neutralizing viral infection. As the convalescent serum is composed of different antibodies, it is expected that fully neutralization should be accomplished by the action of antibodies against these identified epitopes and probably other spatial epitopes. When we were preparing this manuscript, a similar study that used an 18-mer-peptide pool of spike proteins to screen 25 COVID-19 convalescent serums by ELISA. They identified two dominant epitopes that partially overlapped with the peptide S1-55 and S2-13 identified in this study (15). And depletion of the COVID-19 convalescent serums by these two epitopes partially impaired the neutralizing activity of the convalescent serums (15). In these study, in addition to these two peptides, we identified three other dominant epitopes: the epitopes in peptide S1-57 that is close to the peptide S1-55, the peptide S2-47 that is near the membrane proximal region of S and the ectodomain of M (Fig. 1 and 2). Notably, peptide S1-57 competed the neutralizing activity of two serums out of four serums tested (Fig. 5), suggesting the antibodies against this peptide play an import role in neutralizing activity. In contrast, peptide S2-13 didn’t compete the neutralizing activity of the serums tested, which is inconsistent to the previous study (15). The discrepancy might be due to the variation of the serums used.

We noticed two peptides S1-52 that is in the receptor-binding domain and S2-56 that is in the endomain of S highly reacted with SARS-CoV-2 negative serums (Fig. 1), pointing to a risk for using the whole S or RBD as a reagent for diagnosis of SARS-CoV-2 antibodies. The dominant peptides S1-57 and M1 specifically reacted with COVID-19 serums and may provide good candidates for diagnosis reagents.

RBD of coronaviruses spike proteins mediate virus entry and antibodies against RBD potently neutralize viral infection. To our surprise, we detected much less interactions of the peptides in RBD with the serums, except for the peptide S1-52 (Fig. 3). Of the 120 serums, only 5 serums completely neutralized and 7 serums partially neutralized viral infection at the dilution of 1:200. The relative low percentage of neutralizing antibody might be due to the poor recognition of RBD linear epitopes. Alternatively the most of the RBD epitopes are conformational epitopes. We can not rule out the possibility that the non-neutralizing antibodies could sterilize viral infection by other mechanisms such as antibody-dependent cellular cytotoxicity (ADCC) (16).

In summary, we identified the dominant B-cell linear epitopes of SARS-CoV-2 in a COVID-19 cohort. Antibodies elicited by some of these antibodies played an important role in neutralizing viral infection. These epitopes may serve as a vaccine candidate to elicit neutralizing antibodies in most vaccinated people or a specific antigen for SARS-CoV-2 diagnosis.

## Methods

### Ethics statements

This study was approved by the ethics committee of Shanghai public health clinical center, and the procedures were carried out in accordance with approved guidelines. Informed consent was obtained from the subjects. Collection of the COVID-19 convalescent serum samples was approved under the study number YJ-2020-S077-02. The serum samples of patients with symptoms of acute respiratory infections were collected under the study number YJ-2018-S045-02.

### Peptide array

According to a sequence of SARS-CoV-2 (Accession number, MT121215), 20-mer overlapped peptides covering the entire spike (S) region, the ectodomain of membrane (M), the loops between the TMs and representative regions of endomains of M and envelope (E) were synthesized by GL Biochem (Shanghai, China) and purified by HPLC. Peptides were printed onto activated integrated poly (dimethysiloxane) membrane (iPDMS) as described (14).

### Serum screening

Serums were diluted (1:200) and incubated with peptide array. After wash, the peptide arrays were incubated with Horseradish peroxidase (HRP)-conjugated goat anti-human IgG. Dots were visualized by Super Signal Femto Maximum Sensitivity Substrate for Chemiluminescence (Thermo Fischer) and pictures were captured by a cool CCD. The chemiluminescence intensity of each dot was converted to signal-to-noise ratio (SNR) by subtracting the background intensity averaged from the intensity from blank dots. SNR equal or above 2 is considered as positive.

### Cells

Vero E6 cells were maintained in Dulbecco’s modified eagle medium (DMEM)(Hyclone) containing 10% FBS (BI) supplemented with 1% penicillin, and streptomycin (Hyclone).

### Virus and neutralization assay

SARV-CoV-2 virus experiment was carried out in the biosafety level 3 (BSL-3) laboratory in the Shanghai medical college, Fudan University. SARV-CoV-2 virus strain nCoV-SH01 was isolated from a throat swab sample on Vero E6 cells (13), propagated and titrated on Vero E6 cells by a plague assay. For neutralization assay, 5×10^4^ Vero E6 cells were seeded onto 96 well plates. Serums were diluted 1:100 in DMEM supplemented with 2% FBS, incubated with 100 PFU of SARV-CoV-2 virus at 37°C for 1 hours, and then the mixture was added to equal volume of cell medium (the final dilution of serum is 1:200). After infection for 2 hours, cells were washed once with DPBS and added with fresh media. For peptide competing experiment, serums were diluted 1:100 in DMEM supplemented with 2% FBS, incubated with 100 PFU of SARV-CoV-2 virus and indicated peptides at a conerntration of 0.5mg/ml at 37°C for 1 hours, and then the mixture was added to equal volume of cell medium (the final dilution of serum is 1:200 and the final concentration of peptide is 0.25mg/ml).

### Quantitative real-time PCR

Supernatants from infected cells were mixed with Trizol LS (Invitrogen). RNAs were extracted by phenol/chloroform and precipitated by isopropanol. After wash with 70% ethanol, RNAs were dissolved in nuclease-free water and quantified by a Taqman-based real time PCR with One Step PrimeScript RT-PCR kit (Takara) according to the manufacturers’ protocol with 400nM of each primer (sense primer: GAA AAT AGG ACC TGA GCG CAC C, anti-sense primer: TGA CAA TAC AGA TCA TGG TTG CTT TGT) and 200nM of probe (FAM- TAG ACG TGC CAC ATG CTT TTC CAC TGC TTC AGA C -BHQ1). The relative RNA levels were calculated by 2^-ΔCT^ method (17).

### COVID-19 screening

We selected throat swab samples of patients with symptoms of acute respiratory infections that were collected before the COVID-19 outbreak and extracted the nucleic acid. COVID-19 diagnosis was performed by Quantitative real-time PCR as described above.

### Data processing

MegAlign (DNAstar) was used for protein sequence alignment analysis. Transmembrane helices prediction was done by TMHMM program (http://www.cbs.dtu.dk/services/TMHMM-2.0/). Structure data of open state of S was retrieved (PDB, 6vyb) and visualized by UCSF Chimera.

## Data Availability

All data generated or analysed during this study are included in the manuscript and supplementary files.

## Author Contributions

Conceived the study: J Chen, T Ying, Z Yi; conducted the study: K Hu, Y Wang, W Song, R Zhang, Z Yi; clinical sample: Y Ling, X Zhang, H Lu; Data analysis: Z Yi, X Zhang, K Hu, Y Wang; Manuscript draft: Z Yi; Resources: Z Yuan, H LU, Z Yi.

